# Clinical analysis of myocardial injury in highlanders with pulmonary hypertension

**DOI:** 10.1101/2023.05.15.23290019

**Authors:** Maolin Zhao, Qianjin Wu, Wangsheng Duanmu, Junxian Shen, Weixin Yuan, Yingbin Sun, Xu Zhang, Jinbao Zhang, Siyi He

## Abstract

**Background:** High altitude environment can give rise to Myocardial injury (MI) mainly because of hypoxia, where MI with pulmonary hypertension (PH) is one of the severe pathologies. In the present study, we intend to explore clinical characteristics of MI in patients with PH at high altitude and diagnostic value of various myocardial markers.

**Methods:** Consecutive patients at the altitude of 3650m were selected into this retrospective study. Clinical and biochemical data were collected. According to the results of Cardiac troponin I (cTnI), patients were divided into MI group and non-MI group.

**Results:** A total of 231 patients were enrolled in this study. MI occurred in 29 patients (12.6%). We found that body mass index (BMI, *P*=0.045), left ventricular end-diastolic dimension (LVEDD, *P*=0.005), and serum level of creatine kinase-MB (CK-MB, *P*=0.001) in MI group were significantly higher than that in non-MI group. Spearman correlation analysis revealed that cTnI have a significant positive correlation with CK-MB (P=0.000) and LDH (*P* <0.001) instead of aspartate aminotransferase (AST). A receiver operating characteristic (ROC) curve was drawn to demonstrate that CK-MB could significantly predict the occurrence of MI with an area under the curve (AUC) of 0.749 (*P*=0.000), and the level of 3.035 (sensitivity = 59.3%, specificity = 90.5%) was optimal cutoff value.

**Conclusion:** The incidence of MI with PH is high in highlander. As a convenient and efficient marker, CK-MB is closely associated with cTnI and have a predict role in the occurrence of MI with PH under expose to high altitude hypoxia.

## 1. Introduction

The number of people visiting and living in high altitude has increased in recent years. It is estimated that 81.6 million people live at > 2,500 m and 14.4 million people live at ≥ 3,500 m[1]. High altitude is characterized by hypobaric and hypoxia. The heart is highly sensitive to hypoxia and prolonged exposure to altitude-associated chronic hypoxia may cause myocardial injury (MI), which is characterized by varying degrees of elevation of serum myocardial enzymes. High altitude environment tends to cause increased pulmonary circulation resistance, thus formating pulmonary hypertension (PH), which is one of the most common abnormal cardiovascular complications[3]. Hypoxia-induced PH is a major health problem at high-altitude regions of the world. Previous study on Kyrgyz highlanders have reported 14– 20% of the population with PH[4]. Furian et al. [5] suggested that highlanders with PH at high altitude may be at increased risk of cardiovascular mortality and morbidity. Therefore, more attention should be paid on the MI with PH at high altitude.

MI is a severe complication in population exposed to high altitude and may leads to malignant arrhythmias, heart failure, and even sudden death[6]. Our previous study found that the incidence of MI was as high as 33.2% in population transferring from high altitude[7]. Hypoxic and hypobaric environment will cause the damage of the cardiac cells, leading to the elevation of serum myocardial enzymes to varying degrees[8]. Cardiac troponin I (cTnI) and T (cTnT) are expressed almost exclusively in the heart, which are preferred biomarkers of MI with high specificity and sensitivity[9]. Thus, they have been regarded as the gold-standard marker for MI[10]. However, it is difficult to detect cTnI for the reason that the work of instruments is limited at high altitude. Thus, it is important to investigating the other classical MI markers to replace troponin for the prevention and diagnosis of MI at high altitude, such as creatine kinase (CK), lactic dehydrogenase (LDH), and aspartate aminotransferase (AST)[11]. Several studies have used these myocardial enzymes as indicators for the evaluation of MI at high altitude. He et al. [12] found that CK, CK-MB, and LDH levels altered in high-altitude de-acclimatization syndrome after exposure to high altitudes. Zhou et al. [13] suggedted that the cardiac function index was negatively correlated with the levels of creatine kinase-MB (CK-MB) and LDH.

In this study, population exposed to high altitude with PH were included, and comparisons between MI and non-MI were analyzed. We intended to explore the diagnostic value of various myocardial markers for MI in patients with PH at high altitude, thereby providing theoretical basis for early clinical prevention and treatment.

## 2. Materials and Methods

### 2.1 **Study settings**

Consecutive patients admitted to the department of cardiovascular medicine in a general tertiary hospital at the altitude of 3650m were included. Inclusion criteria: (1) Based on ESC/ERS Guidelines for the diagnosis and treatment of pulmonary hypertension[14], all included patients were diagnosed with PH by using echocardiography with the criteria of tricuspid valve regurgitation (TVR) peak velocity > 280 cm/s; (2) adult patients aged >18 years old; (3) living continuously at high altitude (>3000m) more than one year. Exclusion criteria: (1) all kinds of cardiomyopathy, including ischemic, dilated, hypertrophic, and so on; (2) pulmonary embolism; (3) myocarditis; (4) congenital heart disease. According to the fourth universal definition of myocardial infarction[15], patients were diagnosed with MI once the serum level of cTnI was greater than 0.1ng/ml, who were classified into MI group while the others were classified into non-MI group.

### 2.2 Data collection

Basic information of patients was collected through electronic medical record, including age, gender, body mass index (BMI), living altitude, aborigines, and smoke. Echocardiography and electrocardiogram were performed after admission, and then we could get the indexes including TVR velocity, left ventricular end-diastolic dimension (LVEDD) and heart rate (HR). Early morning fasting blood was drawn from all patients after admission for examination to acquire the data about red blood cell count (RBC), hemoglobin (Hb), platelet count, red blood cell distribution width (RDW), platelet distribution width (PDW), alanine transaminase (ALT), AST, total bilirubin, direct bilirubin, albumin, globulin, urea nitrogen, creatinine, uric acid, total cholesterol, triglyceride, high density lipoprotein cholesterin (HDL-C), low density lipoprotein cholesterin (LDL-C), apolipoprotein A1 (ApoA1), apolipoprotein B (ApoB), LDH, CK-MB, prothrombin time (PT), activated partial thromboplastin time (APTT) and D-dimer (D-D).

### 2.3 Statistical analysis

The software SPSS version 25.0 was used to perform statistical analysis, and difference was considered to be significant once P value was lower than 0.05. Normal distribution was examined according to Kolmogorov-Smirnov test. Normally distributed measurement data were expressed as mean value ± standard deviation, and comparisons between groups were analyzed by using t test. Abnormally distributed measurement data were expressed as median value (Q1-Q3), and comparisons between groups were analyzed by using Mann-Whitney U test. Comparisons of enumeration data between groups were analyzed by using chi-square test. Spearman analysis was used to evaluate the correlation between two indexes. Diagnostic accuracy was evaluated by receiver operating characteristic (ROC) curve as assessed by area under the ROC curve (AUC). The optimal cut-off values were determined by using the Youden index.

### 2.4 Patient and public involvement

This study was retrospective, and all patients were not initiative to be enrolled in the present research. Medical data were collected through electronic medical record system. No personal information about the patients were included in this study.

## 3. Results

### 3.1 **Clinical features of MI with PH patients at high altitude**

A total of 231 patients at high altitude with PH were included in the present study, of which 29 patients were diagnosed with MI, accounting for 12.6%. As Table 1 showed, patients in MI group had higher BMI than those in non-MI group [(26.1±4.0) kg/m2 vs. (24.4±4.2) kg/m2, P=0.045]. LVEDD was significantly increased in MI group compared with non-MI group [44(41.5-52.5) mm vs. 42(40-45) mm, P=0.005]. Once MI occurred, the serum level of CK-MB could also significantly upregulate [3.47(2.1-7.6) U/L vs. 1.37(1.1-2.4) U/L, P=0.001].

**Table 1.**
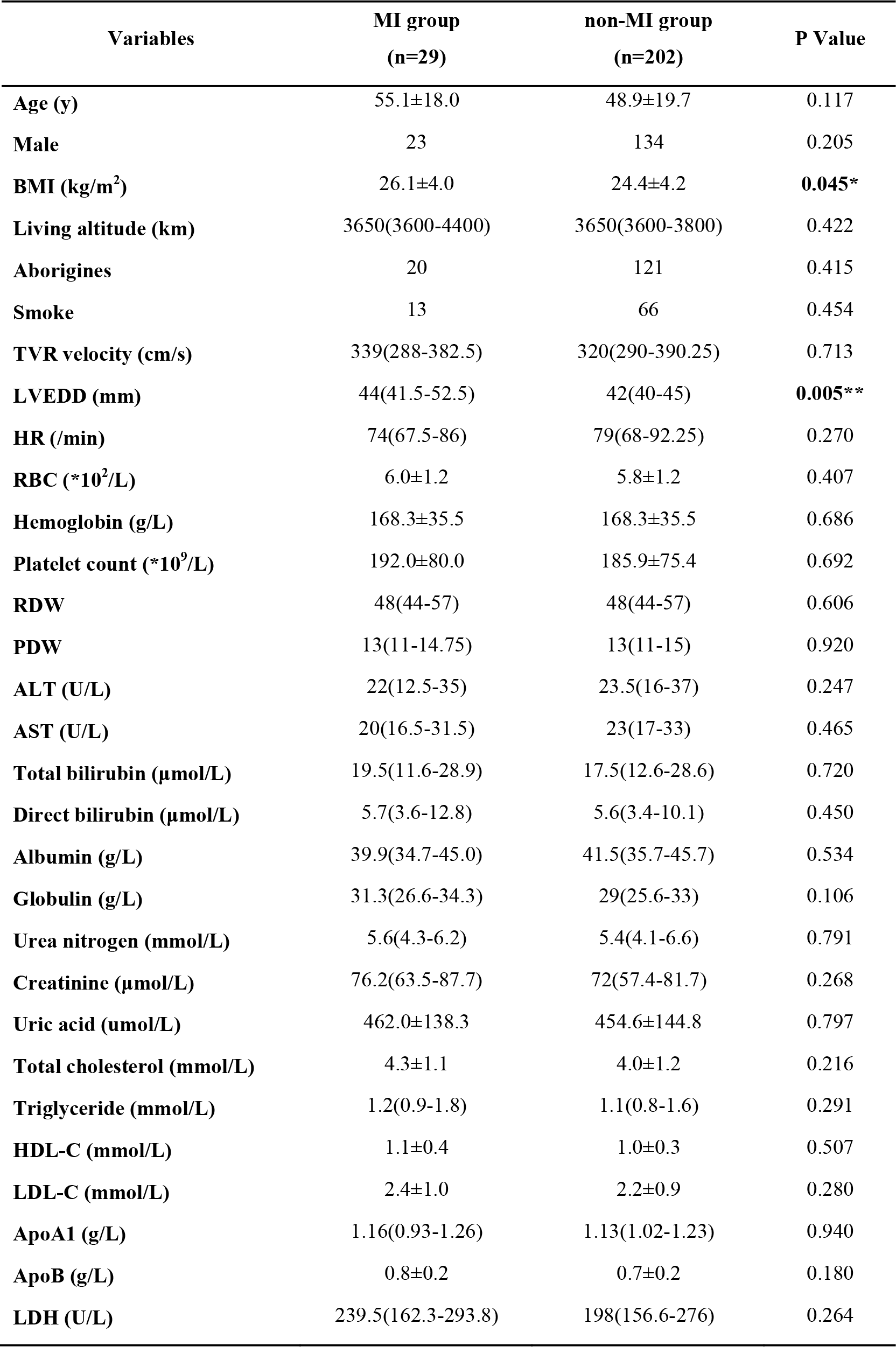
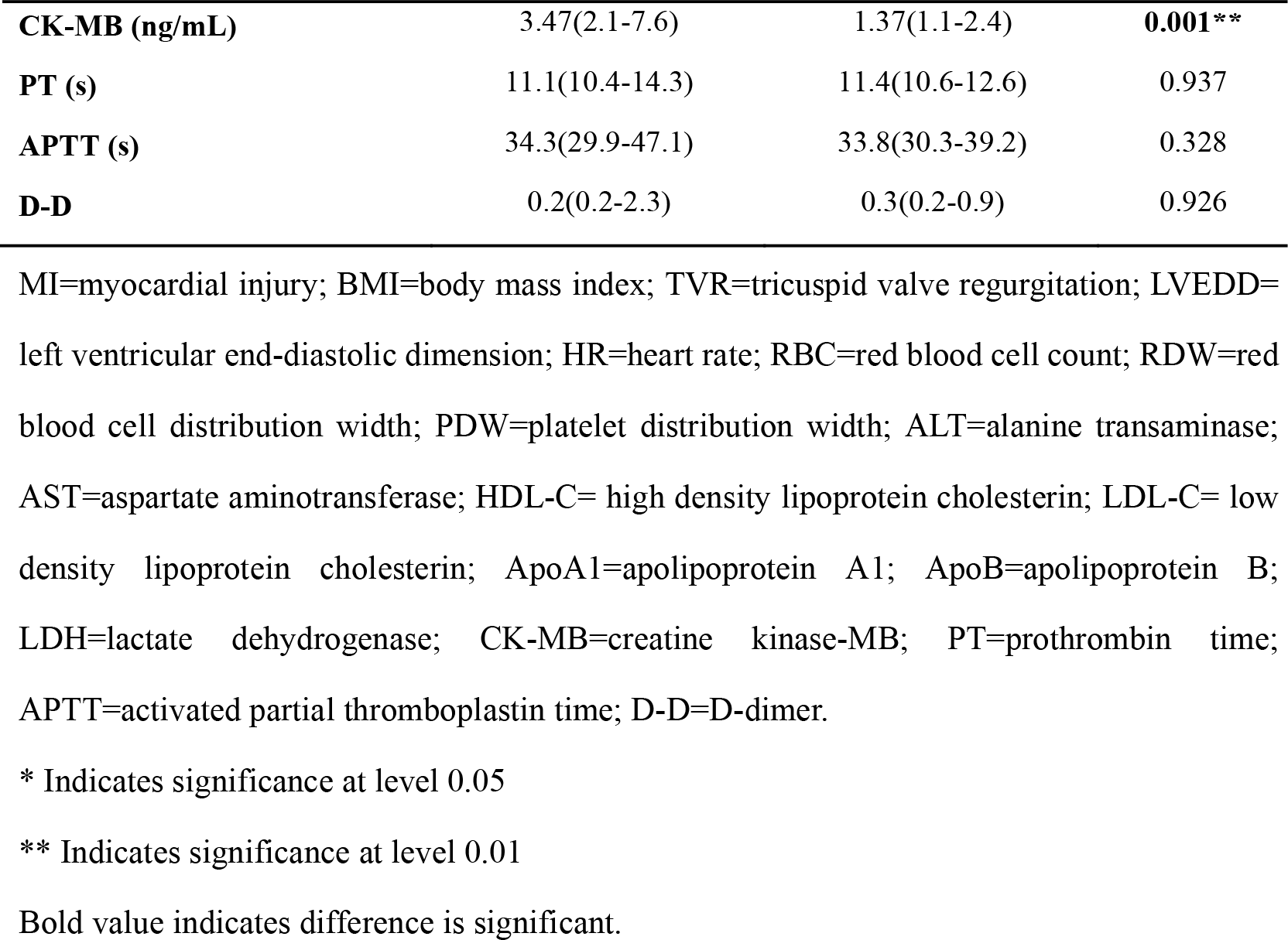
Comparisons of clinical data between MI group and non-MI group.

### 3.2 Correlation between cTnI and other myocardial enzymes

As Table 2 showed, the level of cTnI was positively correlated with the level of CK-MB (r=0.571, P=0.000) and LDH (r=0.212, P=0.001). There was no significant correlation between the level of AST and cTnI.

**Table 2.**
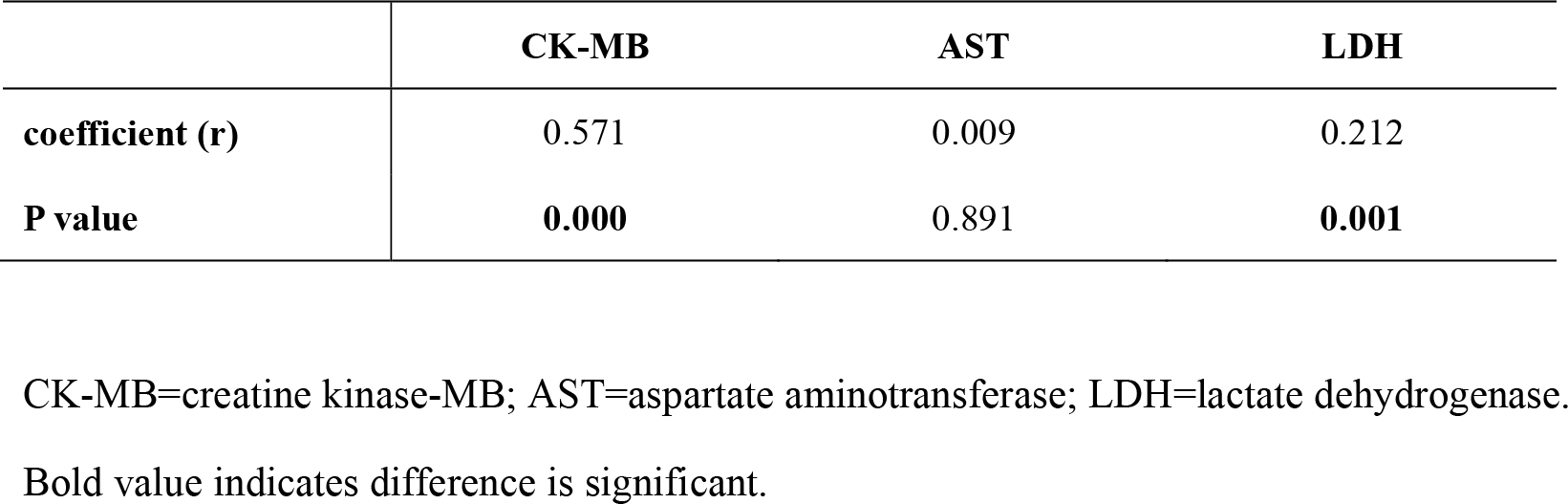
Correlation between cTnI and other myocardial enzymes.

### 3.3 Diagnostic value of myocardial markers

ROC curves were drawn to evaluate the usefulness of classic myocardial markers including CK-MB, AST and LDH in diagnosing MI, as presented in Figure 1. As shown in Table 3, only CK-MB had a significant diagnostic value in MI with AUC of 0.749 at the optimal cut-off value of 3.035 (P=0.000). The sensitivity was 0.593, and the specificity was 0.905. AST and LDH had no diagnostic significance for MI in PH patients at high altitude.

**Figure 1.**
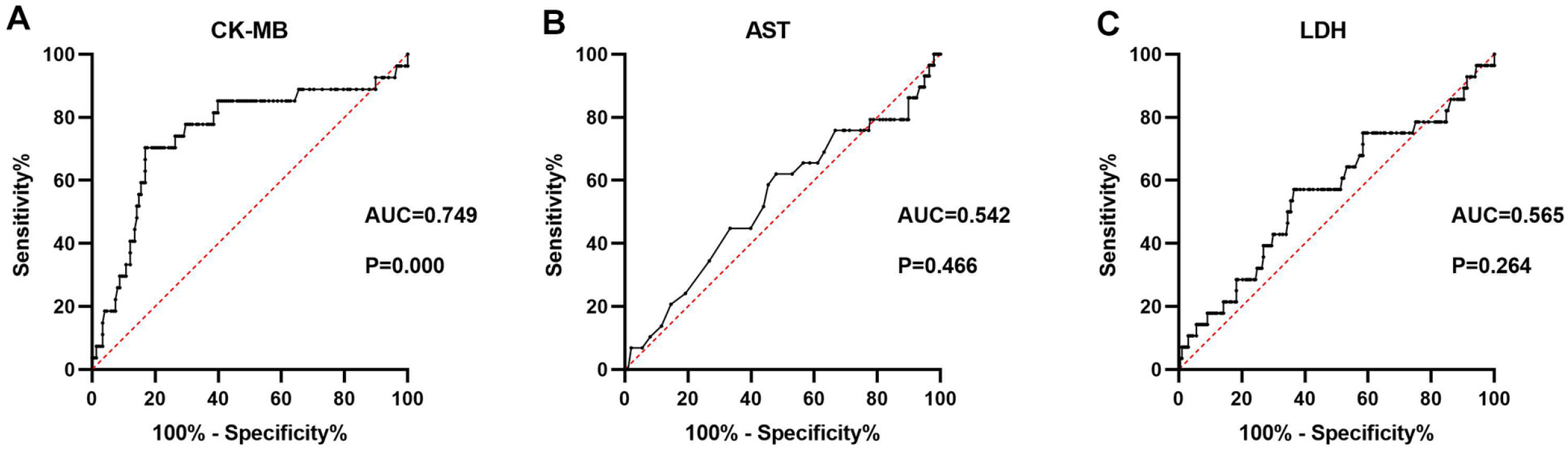
Receiver operating characteristic (ROC) curve for the values of (A) creatine kinase- MB (CK-MB), (B) aspartate aminotransferase (AST) and (C) lactic dehydrogenase (LDH) in diagnosing myocardial injury with pulmonary hypertension.

**Table 3.** Diagnostic value of CK-MB, AST and LDH for MI.

|  | **CK-MB** | **AST** | **LDH** |
| --- | --- | --- | --- |
| **AUC** | 0.749 | 0.542 | 0.565 |
| **Youden index** | 0.498 | 0.141 | 0.206 |
| **Cut-off value** | 3.035 | 22.500 | 233.500 |
| **Sensitivity** | 0.593 | 0.379 | 0.571 |
| **Specificity** | 0.905 | 0.480 | 0.635 |
| **P value** | **0.000** | 0.466 | 0.264 |

## 4. Discussion

Millions of tourists and climbers visit high altitudes annually. Many healthy individuals may suffer from altitude illnesses when visiting to these high regions, including acute mountain sickness (AMS), high altitude cerebral edema and high altitude pulmonary edema (HAPE) and, notably, PH, which can eventually lead to right ventricle hypertrophy and heart failure[16]. Furthermore, increased pulmonary artery pressures, acute hypoxia, increased myocardial work, and increased epinephrine release may cause or exacerbate MI at high altitude[17]. At present, there are few studies focusing on MI with PH at high altitude. In the present research, the prevalence of MI was 12.6%, which lower than our previous study[7, 18]. One possible reason for the lower prevalence was that different diagnostic standards were used. To be specific, due to the limitation of high altitude environment, the previous studies used CK, CK-MB, LDH, and AST to diagnose MI, while cTnI was used in present study. In addition, altitude acclimatization may be another important contributor. This study mainly included population with long-term exposure to the high altitude and aborigines, so adaptive changes may occur in the cardiovascular system, making the heart more tolerant to hypoxia. Adaptation to high-altitude hypoxia increases the cardiac tolerance of highlanders to all the major deleterious consequences of acute hypoxia. Hypoxia inducible factor (HIF) pathway, mitochondrial synthesis pathway, blood oxygen transport pathway, tissue oxygen transport pathway, energy metabolism pathway and other pathways play an important role in the process of myocardial adaptation to high altitude[19]. Evidence suggested that modifications of the HIF pathway is a crucial mechanism in altitude acclimatization. EGLN1 and EPAS1 is encode major components of the HIF transcriptional pathway, which coordinates an organism’s response to hypoxia[20–22]. In a hypoxic environment at high altitude, the hearts initiate transcriptional programs to increase dependence on carbohydrates over fatty acid for ATP production, with altered mitochondrial structure and function that improve metabolic efficiency and reduce ROS generation, as well as beneficial changes in glycogen storage and high energy phosphate system to regulate the balance between energy demand and energy supply when oxygen is limited[23]. In addition, Schweizer et al. [24] found that heart rate is increased to improve the delivery of circulating oxygen at high altitude, and attenuation of catecholamine release improves tissue blood flow during hypoxia. The changes of Hb function, oxygen transport and aerobic capacity under high altitude play a vital role in the process of hypoxic adaptation[25], including the evolution of higher rates of alveolar ventilation and respiratory oxygen uptake[26], higher Hb O2 affinity[27], as well as altered skeletal muscle phenotypes enhance the ability of tissue O2 diffusion and O2 utilization[28, 29].

It is well known that obesity has adverse effects on cardiovascular hemodynamics and cardiac structure and function, thus increasing the incidence of adverse cardiovascular events[30]. Evidence suggests that obesity is a risk factors for AMS and is associated with its development[31]. In present study, MI group had a higher BMI than the non-MI group. Thus, Elevated BMI may be a risk factor for MI expose to high altitude. Previous study demonstrated that among individuals without cardiovascular disease, higher BMI was independently linearly associated with subclinical MI[32]. Obesity leads to myocardial dysfunction by endocrine and inflammatory effects of adipose tissue[33]. However, what’s interesting is that obesity appeared to be associated with a lower risk of death once cardiovascular disease was present[34]. This phenomenon has been called the “obesity para- dox”[35].

In this retrospective research, we also found that LVEDD altered significantly in MI with PH under high-altitude exposure. Left ventricular dilatation is usually the result of multiple pathological changes that occur during myocardial remodeling in progressive heart failure. There is an outward remodeling of the left ventricular myocardium during MI, which results in left ventricular dilatation[36]. In our study, the presence of increased LVEDD in MI group may indicate the heart was going into decompensation phase. Dilated LVEDD was related to left ventricular dysfunction and may be a signal of advanced heart disease[37]. Left ventricular dilatation is an independent predictor of MI[38]. Chen et al. [39] suggested that LVEDD may be used to identify heart fail patients who require more aggressive therapeutic interventions. Left ventricular (LV) enlargement is an independent predictor of cardiovascular events in patients, and the addition of LV volumes to LV diameters increased the predictive value for cardiovascular outcomes [40]. Therefore, screening for LVEDD dilatation in the general population at high altitude may identify a high-risk population for preventive treatment.

Although Troponin has been regarded as the gold-standard biomarker for MI[41], other myocardial enzymes may be helpful in cases where troponin cannot be detected. CK-MB is relatively specific for myocardial tissue, and CK-MB levels are most valuable for the diagnosis of myocardial infarction when skeletal muscle injury or disease is excluded, but the expression of AST and LDH in a wide variety of tissues significantly affects its specificity for MI[42]. In our study, it has been suggested that CK-MB and LDH were closely associated with the expression of cTnI. Furthermore, ROC curve was drawn to demonstrate that CK-MB level could predict the occurrence of MI with an AUC of 0.749 to a limited extent. Similarly, Previous studies have suggested that CK-MB provides a reliable and specific diagnosis with high accuracy in the first few hours of onset of cardiac symptoms[43]. However, AST and LDH were dropped due to lack of specificity in the diagnosis of acute myocardial infarction[44]. Thus, we suggest that CK-MB may be a good diagnostic marker when it is hard to measure of cTnI in highlanders.

## 5. Conclusion

In summary, incidence of MI with PH is high in population at high altitude. BMI, LVEDD and CK-MB altered significantly in MI with PH under high altitude exposure. As a convenient marker, CK-MB is found to be closely associated with cTnI and have an effective diagnostic value in the high-altitude MI with PH. The biggest limitation in our present study is that only patients with PH are included, so whether the findings can be extended to all population at high altitude still needs further study.

## CRediT authorship contribution statement

MZ: Data curation, Formal analysis, Writing-Original Draft; QW: Data curation, Validation, Visualization; WD: Data curation, Writing-Original Draft; JS: Data curation, Validation; WY: Data curation, Validation; YS: Data curation, Validation; XZ: Formal analysis, Writing-Reviewing and Editing; JZ: Supervision, Writing-Reviewing and Editing; SH: Conceptualization, Supervision, Funding acquisition, Writing-Reviewing and Editing.

## Publisher’s note

All claims expressed in this article are solely those of the authors and do not necessarily represent those of their affiliated organizations, or those of the publisher, the editors and the reviewers. Any product that may be evaluated in this article, or claim that may be made by its manufacturer, is not guaranteed or endorsed by the publisher.

## Data Availability

The data are available from the corresponding author on reasonable request.

